# The modified COVID-19 Yorkshire Rehabilitation Scale (C19-YRSm) patient-reported outcome measure for Long Covid or Post-COVID syndrome

**DOI:** 10.1101/2022.03.24.22272892

**Authors:** Manoj Sivan, Nick Preston, Amy Parkin, Sophie Makower, Jeremy Gee, Denise Ross, Rachel Tarrant, Jennifer Davison, Stephen Halpin, Rory J O’Connor, Mike Horton

## Abstract

**Background:** The C19-YRS was the first validated scale reported in the literature for patient assessment and monitoring in Long Covid or Post-COVID syndrome. The 22-item scale contains four subscales measuring symptom severity, functional disability, overall health and additional symptoms.

**Objectives:** This study aimed to modify and refine the scale based on psychometric properties, emerging evidence on additional Long Covid symptoms, and feedback from a working group of patients and healthcare professionals.

**Methods:** Data were collected from 370 patients who completed the C19-YRS scale in a community Long COVID service. The psychometric properties of the Symptom Severity and Functional Disability subscales were assessed using a Rasch Measurement Theory framework, where all individual scale items were assessed for model fit, local dependency, response category functioning and differential item functioning (DIF) by age group and sex. Additionally, the subscales were assessed for targeting, reliability and unidimensionality. The overall health subscale is a single item, and the additional symptoms subscale is not intended to be summed, therefore neither is appropriate for Rasch analyses. Psychometric results and implications were relayed back to the working group for discussion, alongside clinical evidence of emerging and relevant symptoms not covered by the original C19-YRS.

**Results:** Rasch analysis revealed promising psychometric properties of the symptom severity and functional disability subscales, with both displaying good targeting and reliability, although some individual measurement anomalies were noted. The original 0-10 item response category structure did not operate as intended for both the subscales. Post-hoc rescoring suggested that a 4-point response category structure would be more appropriate for both the subscales, and this aligned with patient feedback. This scoring change was implemented, alongside changes in the item composition of the symptom severity and additional symptoms subscales. The functional disability item set, and the overall health single-item subscale remained unchanged.

**Conclusion:** A modified version of the C19-YRS was developed based on a combination of psychometric evidence, clinical relevance of the content and feedback from the working group (comprising patients and healthcare professionals). Future studies including NIHR funded LOCOMOTION study will undertake large-scale, multi-centre validation of the modified C19-YRS.

## INTRODUCTION

Long Covid (LC) is a term coined by patients and refers to persistent symptoms four weeks after contracting COVID-19 illness.^1^ Ongoing symptomatic COVID-19 and Post-COVID Syndrome (PCS) are the scientific terms for symptoms 4-12 weeks and >12 weeks after the illness respectively.^2^ LC affects more than 2 million individuals in the UK alone and more than 50 million cases worldwide.^3^ More than 200 symptoms across 10 organ systems have been reported with most common symptoms being breathlessness, fatigue, palpitations, dizziness, pain, brain fog (cognitive problems), anxiety, depression, post-traumatic stress, skin rash and allergic reactions.^4^ It can be a remitting and relapsing condition with a protracted course causing significant distress and disability in some individuals.^5^

A multidisciplinary team (MDT) of rehabilitation professionals working with patients recovering from COVID-19 during the first wave of the pandemic developed the original version of C19-YRS.^6-8^ The content was based on staff experience of managing these patients, knowledge from our systematic review of previous outbreaks and feedback on the scale from patients and healthcare professionals.^7-9^ The content was decided using a consensus method and the scale was kept balanced in terms of questions spanning all aspects of the 2001 WHO International Classification of Functioning, Disability and Health (ICF) framework.^10^ The content validity of the scale was supported by studies^11, 12^ using the scale which revealed symptoms and functional problems similar to other LC studies in the literature.^13, 14^

C-19 YRS was the first validated scale reported in the literature to capture LC symptoms and grade the severity of symptoms and functional disability in LC. The use of the scale has been also been recommended in the NHS England Clinical Guidance for LC services and NICE rapid guidelines. ^2, 15^ The scale has been translated in numerous international languages and currently used in many LC studies worldwide. There is also a digital format of the scale available where the patient completes the questionnaire on a smartphone application and the clinicians accesses the results on a web portal and both use the system to monitor progress and response to ongoing treatments for LC. ^8^

The original C19-YRS is a 22-item patient-reported outcome measure (PROM), with each item rated on a 0-10 numerical rating scale, where 0 represents symptom not present and 10 represents symptom being extremely severe or life disturbing. The C19-YRS is broken down into four subscales concerned with the severity of patients’ key symptoms, functional limitations, overall health and additional symptoms. Pre-COVID scores are also captured for comparison.^8^ Q 1-10 form the symptom severity subscale (score 0-100), Q11-15 the functional disability subscale (0-50), Q16 is the overall health score (0-10) and Q 17-22 the additional symptoms subscale (0-60).^16^ The classical psychometric analysis of the C19-YRS in a sample of 188 LC patients showed good data quality, satisfactory scaling and targeting and high internal consistency (Cronbach’s alpha = 0.891), with good reliability of individual subscales.^16^ Some items were identified as having poor scaling assumptions and targeting such as swallowing, incontinence, fever and skin rash, and it was identified that the contribution of these items to the overall measurement properties of the scale was limited.^16^

Although the classical psychometric analysis of the C19-YRS was promising, a further analysis using modern psychometric approaches (Rasch analysis) was included as part of the C19-YRS development plan. The Rasch model^17^ is a unidimensional measurement model that satisfies the assumptions of fundamental measurement,^18, 19^ meaning it provides a measurement template against which scales can be tested. Rasch Measurement Theory (RMT) therefore provides a way to assess the validity of multi-item latent scales where the items are summed together to form an overall total score. RMT provides a unified framework for several aspects of internal construct validity to be assessed, highlighting measurement anomalies within an item set. It should be emphasised that this C19-YRS development phase was intended to identify any particular measurement issues that could help to guide towards a modified version of the C19-YRS that would be psychometrically robust.

Since the development of the original C19-YRS scale early on in the first wave of the SARS-CoV-2 pandemic, important symptoms, such as post-exertional malaise have been identified as clinically important in management of Long Covid. Such symptoms, particularly those identified as important by patients and healthcare professionals need to be considered for inclusion in the modified version of the scale.

The aim of this study was therefore to test the psychometric properties of the scale based on the Rasch model, and to create a modified version of the C19-YRS which optimises the measurement characteristics of the scale, whilst incorporating important insights from patients and healthcare professionals.

## METHODS

### Study design

This was a prospective observational study involving Long Covid patients attending a community based Long Covid service within one of the UK’s largest metropolitan areas, serving a population of approximately 850,000 people. Patients were referred by their General Practitioner (GP) or community therapy teams to the service and they completed the original C19-YRS questionnaire as part of initial triage. Clinicians and patients’ family members or carers were permitted to help complete the responses. On return of the C19YRS, the clinician researchers transferred the data from each completed C19-YRS to an Excel spreadsheet. The data were fully anonymised at the point of statistical team input. A favourable ethical opinion was received from the University of Leeds School of Medicine Research Ethics Committee in January 2021 (reference MREC 20-041 - Secondary analysis of C19-YRS (COVID-19 Yorkshire Rehabilitation Scale) and approved by Leeds Community Healthcare Research and Innovation department.

### Rasch analysis

Rasch analysis was completed with RUMM2030 software,^20^ and carried out separately for the symptom severity subscale (10 items) and the functional disability subscale (5 items). The overall health score is comprised of a single item, which is treated independently from the other subscales, and is therefore inappropriate for Rasch analysis. The additional symptoms subscale was not assessed, as these items provide supplementary information to the clinical staff, rather than contributing to the symptom severity subscale.

A number of scale and item tests of fit were carried out, and these are all described in more detail elsewhere.^21^ All items were assessed for individual fit to the Rasch model, relative to the subscale item set, to test whether each item was contributing to the same underlying construct; misfit was indicated where items were significant at a Bonferroni-adjusted chi-square p-value, or where standardised (z-score) fit-residuals fall outside ±2.5. Tests of local dependency (LD) were carried out to determine whether the response to any item has a direct impact on the response to any other item in the subscale; LD was indicated using a Q3 criterion cut point of 0.2 above average residual correlation.^22^ Response category functioning was assessed to determine whether the response structure of the items was working as intended. For each item, a functional 0-10 response category structure would be indicated by sequential response thresholds (the crossover points between adjacent response categories) on the underlying logit scale. Item bias was assessed through uniform and non-uniform differential item functioning (DIF) testing by sex and age group; with significant DIF indicated at a Bonferroni-adjusted ANOVA p-value. Scale targeting was assessed graphically through the relative distribution of item and person locations. Unidimensionality was assessed through a series of t-tests,^23^ with multidimensionality indicated where independent subsets of items deliver significantly different person estimates, and the lower bound 95% CI percentage of significantly different t-tests is > 5%.

### Working group

A working group comprising five individuals with LC, one dietitian, one psychologist, four physiotherapists, two occupational therapists, two rehabilitation physicians, two researchers and a psychometrician provided feedback on proposed amendments to the scale. The emphasis remained on keeping the scale as brief and comprehensive as possible, without placing undue burden on the respondent.

## RESULTS

### Sample

Data was collected from 370 patients who completed the C19-YRS scale in a community Long Covid service. Key demographics are presented in Table 1.

**Table 1.** Demographics of participants.

|  | **All\*** | **Non hospitalised** | **Hospitalised** |
| --- | --- | --- | --- |
|  | (n=370) | (n=304) | (n=66) |
| Female (%) | 237 (64%) | 208 (68%) | 29 (44%) |
| Mean age (years)(SD) | 47 (14) | 46 (13) | 53 (14) |
| Mean weight (kg)(SD) | 82 (22) | 80 (21) | 93 (22) |
| Mean BMI (kg/m<sup>2</sup>)(SD) | 29 (8) | 28 (8) | 32 (7) |
| Ethnicity (%) |  |  |  |
| *White* | 307 (84%) | 256 (86%) | 51 (78%) |
| *Black* | 10 (3%) | 6 (2%) | 6 (3%) |
| *Asian* | 40 (11%) | 30 (10%) | 10 (15%) |
| *Mixed/Other* | 7 (2%) | 7 (2%) | 0 (0%) |
| Smoking status (%) |  |  |  |
| *Never smoked* | 235 (65%) | 199 (67%) | 36 (55%) |
| *Current smoker* | 24 (7%) | 22 (7%) | 2 (3%) |
| *Ex-smoker* | 105 (29%) | 78 (26%) | 27 (42%) |
| Employment status (%) |  |  |  |
| *Still employed/student* | 176 (49%) | 155 (52%) | 21 (33%) |
| *Still retired/homemaker/unemployed* | 41 (11%) | 34 (11%) | 7 (11%) |
| *Reduced hours* | 48 (13%) | 44 (15%) | 4 (6%) |
| *Sick leave* | 77 (21%) | 48 (16%) | 29 (45%) |
| *Stopped work* | 19 (5%) | 16 (5%) | 3 (5%) |
| Positive COVID-19 test (%) | 228 (62%) | 182 (60%) | 46 (70%) |
| Admitted to hospital (%) | 66 (18%) | 0 (0%) | 66 (100%) |
| Median duration of symptoms (days)(IQR) | 211 (143, 353) | 223 (150, 355) | 159 (129, 288) |

\* Where numbers do not total 370, this is due to missing data

### Rasch Analysis

#### Symptom Scale

Initially, a total of 12 items were entered into the Rasch analysis, as the ‘breathlessness’ section is made up of three separate items: breathlessness at rest; breathlessness on dressing yourself; and breathlessness on walking up a flight of stairs.

These three breathlessness items were immediately identified as displaying a large degree of dependency (pairwise Q3 value 0.57 between breathlessness 1 and breathlessness 2; pairwise Q3 value 0.53 between breathlessness 2 and breathlessness 3; Q3 criterion value indicating dependency = 0.12). Although this finding makes complete conceptual sense, it also means that the separate items should not all be included in contributing to the total score of the symptom severity scale. The breathlessness section was therefore reconfigured so that only the maximum score observed across the three items was used, resulting in a single maximum breathlessness item.

Initial Rasch analysis of the Symptom Severity scale (10 items, including single maximum breathlessness item) looked promising, but revealed certain measurement issues with the item set. Overall scale fit statistics are presented in Table 2. At this point, three items displayed misfit on the chi-square statistic (fatigue, continence, anxiety), with the continence item displaying the largest degree of misfit. Additionally, four pairwise dependencies were identified. Listed in order of magnitude (Q3 criterion=0.10), these were between: anxiety & depression (Q3=0.38); fatigue & cognition (Q3=0.22); anxiety & post-traumatic stress (Q3=0.16); cough & swallowing (Q3=0.14). However, the most substantial issue at this point appeared to be the functioning of the response categories, where all items displayed reverse thresholds. It was apparent that a 0-10 response structure was inappropriate for this item set, as a logical progression of ordered response thresholds was not observed for any of the items (Figure 1). The extent of the disordering was variable depending on the nature and content of the item, with the continence and post-traumatic stress items particularly unsuited to this response structure.

**Table 2.** Rasch Analysis Summary Statistics of SIDECAR scales.

|  |  |  |  | Item Fit Residual |  | Person Fit Residual |  | Overall Chi Square Interaction |  |  |  |  | Unidimensionality T-Tests (CI) |  |
| --- | --- | --- | --- | --- | --- | --- | --- | --- | --- | --- | --- | --- | --- | --- |
|  | Analysis | Number of items | valid n (no. of extremes) | Mean | SD | Mean | SD | Value | df | p | PSI | Alpha | proportion significant | CI |
| Symptom Severity Subscale | Initial | 10 | 368 (1) | 0.31 | 1.36 | -0.20 | 0.99 | 97.6 | 50 | <0.001 | 0.80 | 0.82 | 0.106 | 0.084-0.129 |
|  | Rescored | 10 | 368 (1) | -0.05 | 1.33 | -0.24 | 1.03 | 78.5 | 50 | 0.006 | 0.78 | 0.80 | 0.071 | 0.049-0.093 |
|  | Minus Anxiety | 9 | 368 (1) | 0.02 | 0.90 | -0.24 | 0.96 | 47.8 | 45 | 0.360 | 0.74 | 0.76 | 0.06 | 0.038-0.082 |
|  | Minus Depression | 9 | 368 (1) | -0.05 | 1.06 | -0.26 | 1.00 | 61.2 | 45 | 0.050 | 0.76 | 0.77 | 0.06 | 0.038-0.082 |
| Functional Disability Subscale | Initial | 5 | 350 (18) | -0.08 | 1.77 | -0.34 | 0.99 | 48.9 | 25 | 0.003 | 0.75 | 0.80 | 0.026* | 0.003-0.049 |
|  | Rescored | 5 | 349 (19) | 0.20 | 1.37 | -0.31 | 0.99 | 42.9 | 25 | 0.014 | 0.72 | 0.77 | 0.026** | 0.003-0.049 |
| Target values |  |  |  | 0 | 1 | 0 | 1 | non-significant |  |  | >0.7 | >0.7 | Lower CI < 0.05 |  |

SD = Standard deviation
df = Degrees of freedom
PSI = Person separation index
CI = Confidence intervals
\* = Limited power in unidimensionality t-test
\*\* = Low power in unidimensionality t-test
extremes = people scoring either maximally or minimally across the complete item set

**Figure 1.**
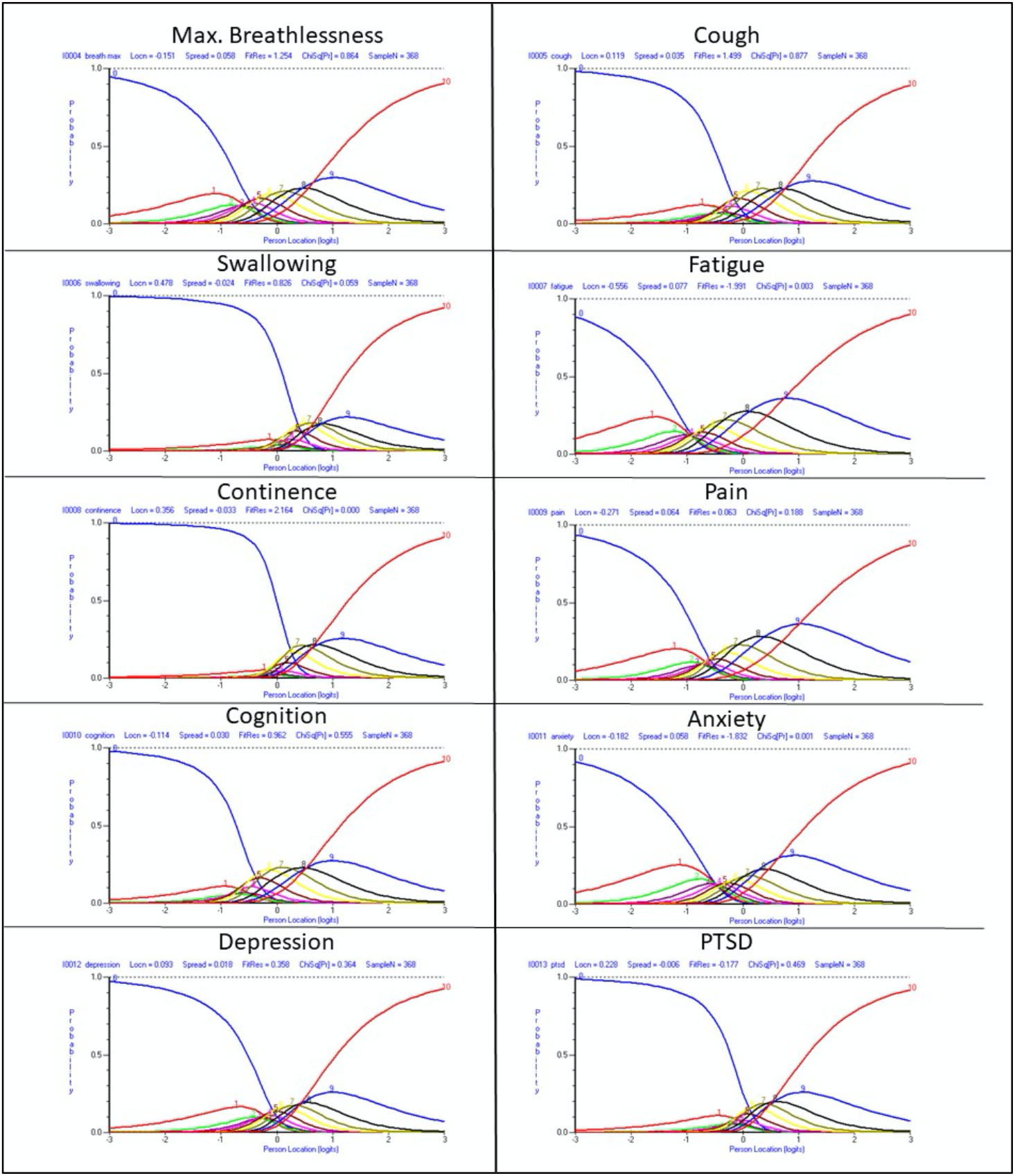
Response category probability curves for each item of the original C19-YRS symptom severity subscale, with 0-10 response structure.

### Rescoring

The inappropriate response structure was discussed within the working group, where it was decided that four response options would seem a reasonable alternative, striking a balance between the number of measurement points and the amount of conceptually different, distinguishable response categories. Various post-hoc rescore options were tested, with the most appropriate 4-response alternative applied across all items appearing to be: 0 (no problem); 1-5 (mild problem/ does not affect daily life); 6-8 (moderate problem/ affects daily life to a certain extent); 9-10 (severe problem/ affects all aspects of daily life/ life-disturbing). It should be noted that this scoring structure was applied post-hoc to the 0-10 scoring system, and that this rescoring is only implied, as respondents have not yet been presented with this 4-category response structure. This resulted in improved threshold ordering across all items, although the swallowing and continence items still displayed slight disordering (Fig 2).

**Figure 2.**
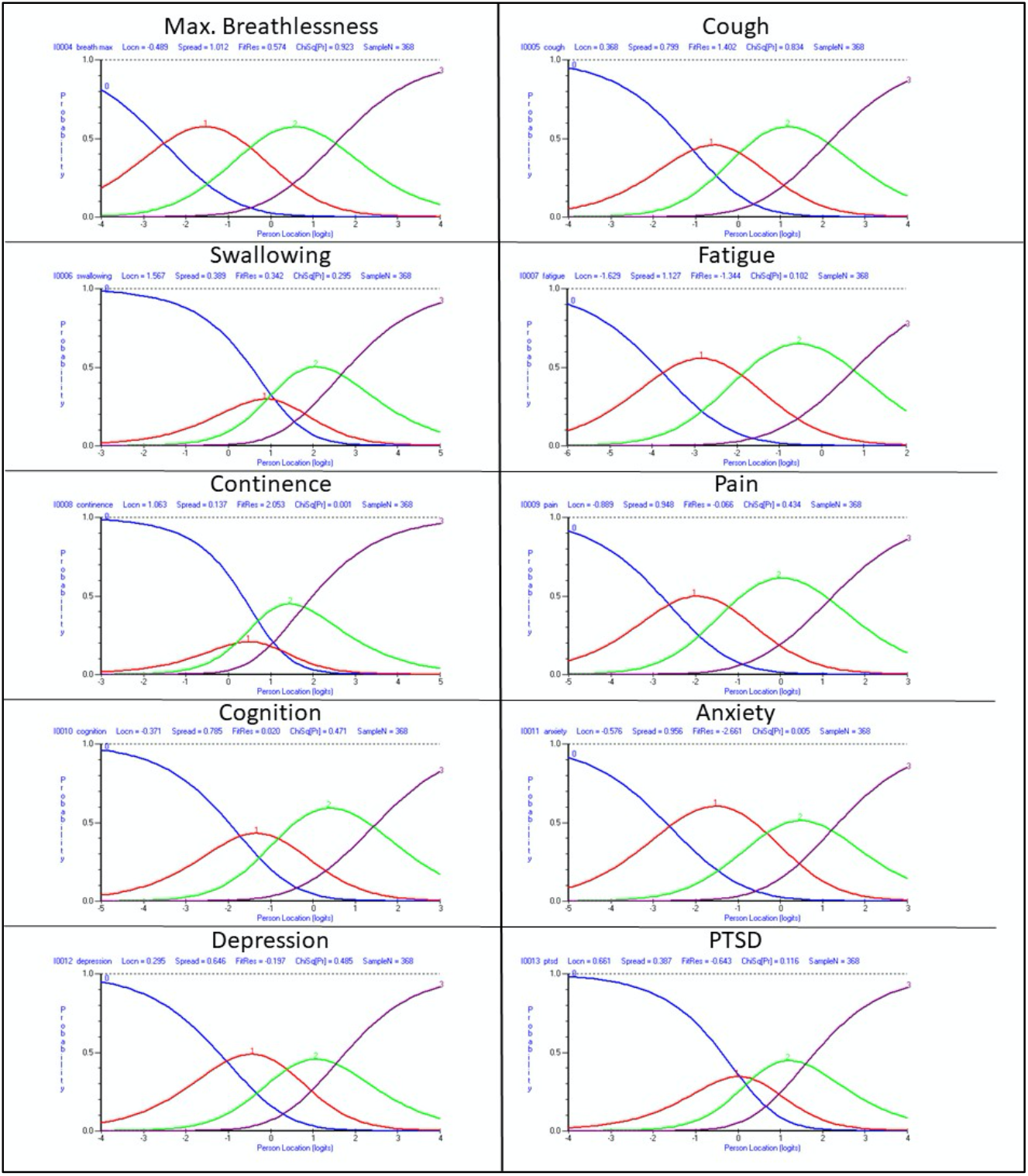
Response category probability curves for each item of the symptom severity subscale, with rescored (implied) four-point (0-3) response structure of the modified C19-YRS.

Overall scale fit statistics following rescoring are presented in Table 2. At this point, two items still displayed misfit on the chi-square statistic (continence, anxiety), with the anxiety item also displaying a fit residual of -2.66. The rescoring had little effect on the pairwise dependencies, which remained present as previously reported. Additionally, there were no differential item functioning (DIF) by sex, age group, or BMI group, and the scale-sample targeting was good (see Figure 3).

**Figure 3.**
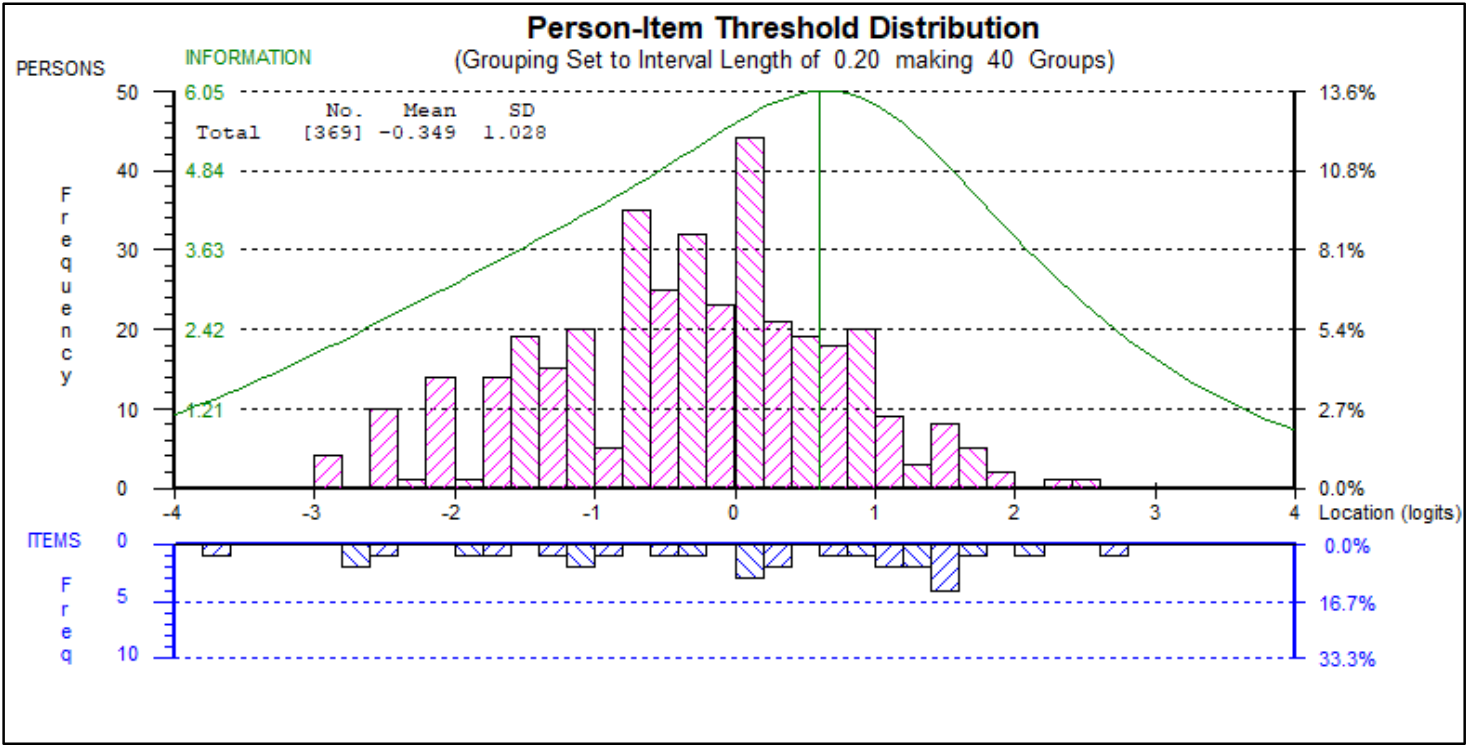
Scale-Sample targeting of Symptom Severity Scale.

Also, although it was not the intention of the study to determine this, distributional differences between demographic groups were observed, with mean score differences by sex (females more severely affected than males, p=0.02), age group (people aged 50+ more severely affected than those below 50, p<0.01), and BMI group (underweight group more severely affected than overweight, who are more severely affected than healthy weight, p<0.001). Further exploratory procedures suggested that the apparent dependency was impacting on the overall fit of the scale, as removal of either the depression item or the anxiety item results in a well-fitting, unidimensional scale (see Table 2).

#### Functional disability scale

Initial Rasch analysis of the functional disability scale (5 items) again looked promising but revealed certain measurement issues with the item set. Overall scale fit statistics are presented in Table 2. At this point, only one item was borderline misfitting on the Chi-square statistic (ADL). Additionally, a pairwise dependency was observed between mobility & personal care (Q3=0.06; Q3 criterion=-0.03). Again, as with the symptom severity scale, the most substantial issue appeared to be the functioning of the response categories, where all items except activities of daily living (ADL) displayed reverse thresholds (not presented), and it is again apparent that a 0-10 response structure is inappropriate for this item set. Items were again rescored in the same 0-4 manner as the symptom items. The overall scale fit statistics following rescoring are presented in Table 2.

At this point, one item still displayed misfit on the chi-square statistic (personal care), and the previously observed pairwise dependency between mobility and personal care was still present. There was no differential item functioning (DIF) by sex or BMI group, although the mobility item does display slight DIF-by-age. Again, the scale-sample targeting was good (see Figure 4).

**Figure 4.**
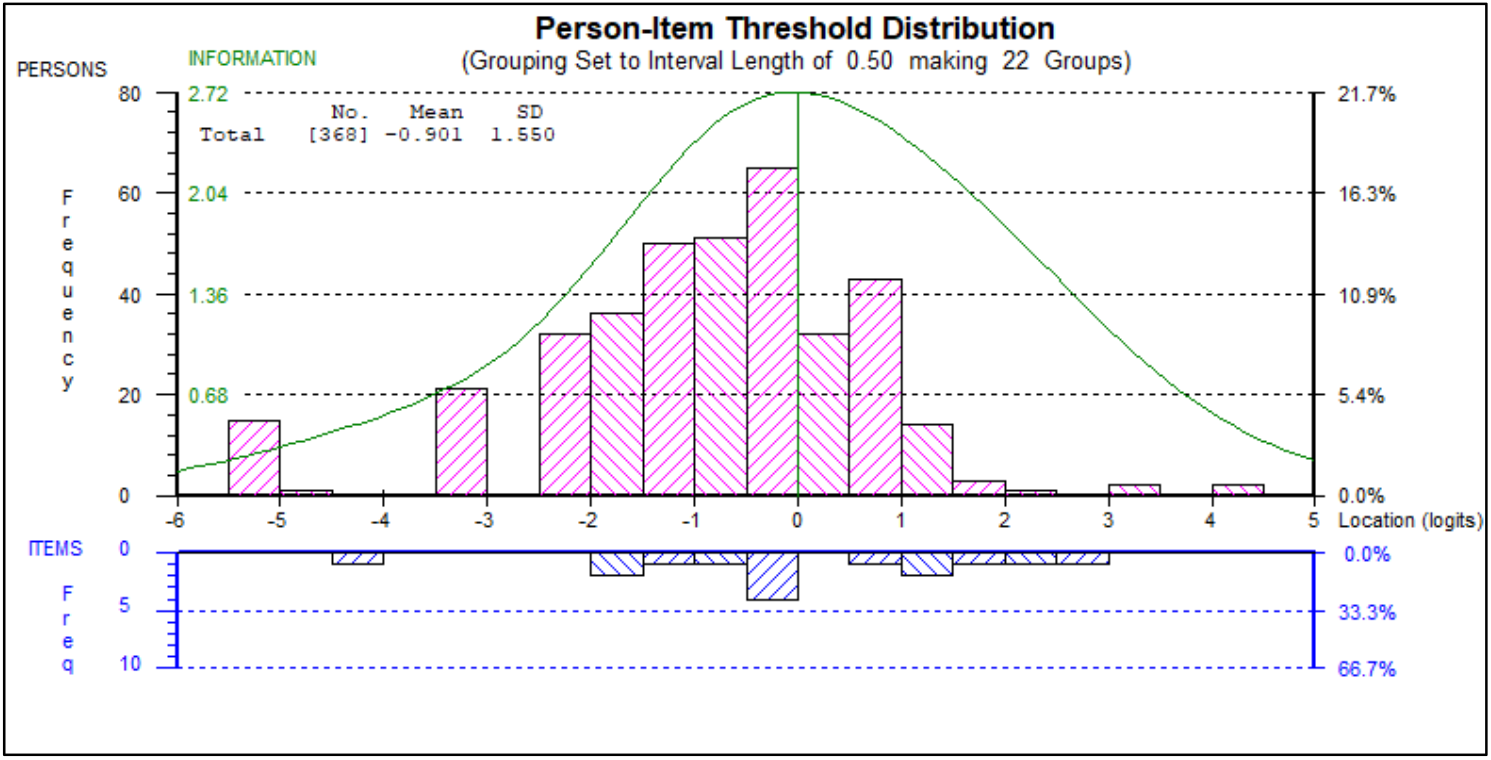
Scale-sample targeting of the functional disability scale.

As with the symptom severity scale, distributional differences between demographic groups were again observed, with mean score differences by sex (females more severely affected than males, p=0.02), age group (people aged 50+ more severely affected than those below 50, p<0.01), and BMI group (underweight group more severely affected than overweight, who are more severely affected than healthy weight, p<0.05).

### Working group

The potential psychometric issues and strengths of the C19-YRS were acknowledged by the working group, and the suggested re-scoring structure was supported across both the symptom severity and functional disability subscales. Additionally, some further items were added to this scale following their emergence as important during clinical presentation and evolving literature (such as post-exertional malaise and altered taste and smell sensation). The apparent dependency of the anxiety and depression items was taken into account by including anxiety/mood as a singular contributing item. As the continence item remained problematic in terms of its fit and response structure, this was moved into the additional symptoms subscale, where a binary response structure was utilised. The final list of main symptoms included in the Modified C19-YRS symptom severity subscale was extended to 10, including breathlessness, cough/ voice, smell/ taste, fatigue, pain/discomfort, cognition, palpitations/dizziness, anxiety/ mood/ post-traumatic stress, sleep, and post-exertional malaise.

The items included in the functional disability subscale remained the same, including communication, mobility, personal care, activities of daily living, and social role. The additional symptoms that are included as a checklist in the Modified C19-YRS are fever, skin rash, allergies, hair loss, eye changes, bruising/ bleeding, visual changes, swallow, balance, weakness, tinnitus, nausea, dry mouth/ ulcers, acid reflux, appetite changes, weight changes, bladder/ bowel symptoms, menstrual cycle changes, sleep apnoea and thoughts of self-harm.

The overall health single-item subscale was retained in its original 0-10 response structure. Additional information regarding family/carers views and vocational aspects were also retained in the modified C19-YRS as they are in the original scale version of the scale. Table 3 lists the key changes in the modified version of the scale along with the reasons for the changes.

**Table 3.** Summary of changes made to the modified C19-YRS (compared to the original C19-YRS)

|  | **Changes made in modified C19-YRS** | **Reason for change** |
| --- | --- | --- |
| **Q1-15** | Response categories changed from 11 to 4 for each of the items of the symptom severity subscale and functional disability subscale | Rasch analysis suggested disordered thresholds for these items (Fig 1) that improved thresholds post-hoc with rescoring (Fig 2) |
| **Q1-10** | Provided the four response categories to each of the symptoms within each single item | Working group suggested it would be easier for respondents to rate each symptom rather than rating only the worst symptom (in the original scale). This change would also help those struggling with brain fog to understand and respond to the question. |
| **Q4** | Capturing altered smell and taste | Working group highlighted the importance of this symptom and emerging evidence on rehabilitation strategies that can be used for these symptoms |
| **Q7** | Palpitation and dizziness introduced as a core symptom | Working group suggested that dysautonomia has emerged as one of core mechanisms linked to many of the Long Covid symptoms. |
| **Q8** | Included post-exertional malaise as a core symptom | Working group and emerging literature recognised this as one of the characteristic features of Long Covid which explains the fluctuating nature of the condition |
| **Q9** | Merged anxiety, mood and post-traumatic stress in one single item | Rasch analysis showed the local dependence of these items when scored separately (as in the original scale) |
| **Q10** | Sleep introduced as a core symptom | Working group suggested to introduce this as one of the key symptoms that characterises Long Covid and was closely related to fatigue and other symptoms |
| **Other symptoms** | Moving swallowing, continence and suicidal idea items to this section | Rasch analysis and working group suggested these symptoms worked more in a dichotomous fashion rather than graded severity of symptom severity scale. Such symptoms with dichotomous responses were placed in the other symptoms section. |
| **Other symptoms** | Introduction of new symptoms: allergy, hair loss, skin sensation, dry/ red eyes, swelling of limbs, bruising/ bleeding, visual changes, tinnitus, nausea, acid reflex, appetite, weight changes, sleep apnoea and changes in menstrual cycles or flow | Working group and emerging evidence suggested even though these are not present in all patients they need capturing as these symptoms can be cause of concern to patients and need addressing by clinicians |

## DISCUSSION

The modified C19-YRS captures the severity of the main persistent symptoms and functional disability in individuals with Long Covid or Post-COVID syndrome. Rasch analysis of the original scale has led to an amendment in the response structure from a 0-10 numeric rating scale to a 0-4 response scale. Additional symptoms subscale includes those with a binary response (yes/no). Symptoms that have been added to the original scale reflect inclusion from the evolving literature and feedback from patients and healthcare professionals based on their understanding of the condition and its impact on health.

The new response category structure will be psychometrically beneficial and is more intuitive for patients, with more distinct response categories. Despite the reduction in response categories, there is also very little reduction in the internal-consistency or reliability of the subscales. The improved response structure may enhance monitoring of the condition at different time points and capture the impact of interventions used in the management of the condition. However, it should be noted that although the amended response structure appeared optimal within this study, it is based on post-hoc collapsing and the operation of the response structure needs to be tested prospectively.

The digital format of the scale (available on ELAROS smartphone application) allows users to track their condition in time and provides them with a visual quantitative assessment of improvement or deterioration of LC which is crucial in the management given less frequent human contact during the pandemic. Clinicians are able to monitor the patient’s progress using the web-based clinical portal of the ELAROS system. Healthcare services can evaluate their treatment programmes using the digital system. National and international comparison of LC data (using the paper or digital format of the scale) can be undertaken while assessing the influence of individual demographics and illness characteristics on LC symptoms.

The World Health Organisation (WHO)’s International Classification of Functioning, Disability and Health (ICF) provides us with a framework to understand the relationship between different aspects of any health condition.^6^ The domains covered by the modified C19-YRS when mapped to the components of ICF (Figure 5) shows that there is satisfactory capture of all the components (body functions and structures, activities, participation, environmental factors and personal factors) making it suitable for a comprehensive biopsychosocial assessment.

**Fig 5.**
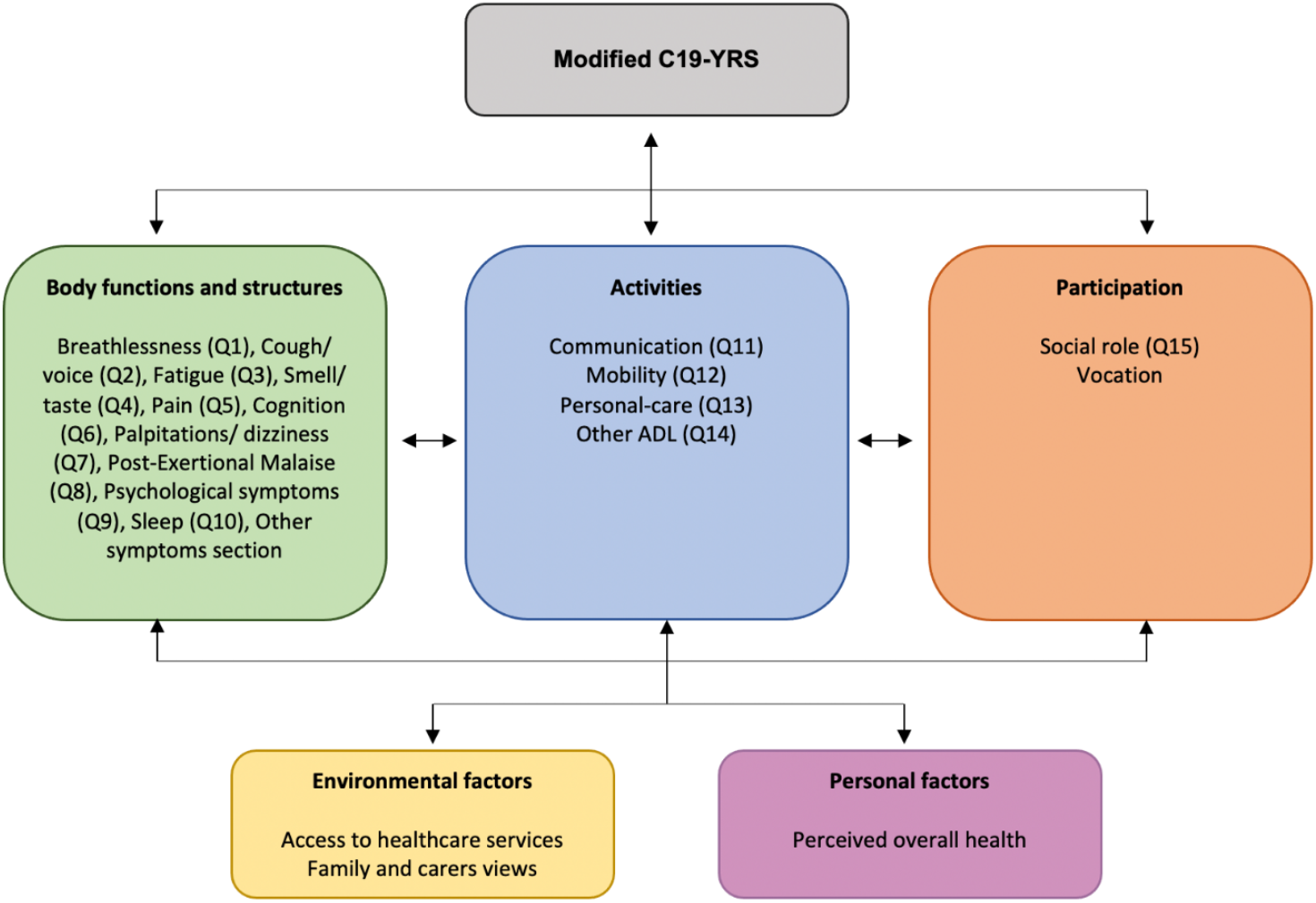
Mapping of the modified C19-YRS tool onto the WHO ICF framework.

Our future work with the scale will involve further evaluation of psychometric properties and validation of the modified C19-YRS in the Long Covid population. The NIHR-funded project Long Covid Multidisciplinary Consortium for Optimising treatments and services acrOss the NHS (LOCOMOTION) is a platform of >5000 patients in the UK whose symptoms and functional limitations will be captured using the modified C19-YRS at regular 3-monthly intervals.^24^ We will have the opportunity to assess the construct and criterion validity of the scale, responsiveness and ability to monitor effect of interventions, along with picking up the natural daily and weekly fluctuations of the condition. This can also estimate how effectively the measure captures differences between individuals, and changes over time within the individual. The floor and ceiling effects of the measure will be assessed to establish the active measurement range of the scale, and we will estimate how effectively the measure captures small differences between individuals at either end of the clinical spectrum of the condition. We will also evaluate the respondent burden of completing the measure within the population, and we will assess the use of digital tools, which can be challenging in certain cohorts (such as those with cognitive problems and those who do not use smartphones). The scale will undergo further Rasch analysis to validate the scale and determine its validity as an outcome measure in LC. Additionally, when the assumptions of the Rasch model are satisfied, it is possible to transform the ordinal-level scale raw scores to an interval-level score, due to the sufficiency of the raw score.^25^ This was not the aim of the current project, but a large-scale validation project of the modified C19-YRS would allow for the creation of a stable interval-level transformation table to be created.

The modified C19-YRS has an advantage over individual symptom-specific measures currently being used in Long Covid studies in that it is comprehensive in covering most symptoms, less burdensome and condition specific (rather than adopting measures that have been developed for other conditions).^26^ There is also an opportunity to explore whether C19-YRS could be developed to a preference-based measure and undertake an economic evaluation of resource use and QALY analysis. The findings of this further research are likely to influence local policy, commissioning and service delivery that is needed to manage the growing number of Long Covid cases worldwide.

## CONCLUSIONS

In summary, a modified C19-YRS has been developed to capture the common symptoms, functional disability and overall health, assessing problems across the multiple body systems affected in Long Covid, and cover all aspects of the WHO ICF framework. The scale allows patients and health care staff to monitor these aspects over the course of the condition, potentially capture Long Covid fluctuations and assess the impact of rehabilitation interventions in the condition.

## Supporting information

Modified C19-YRS

## Data Availability

All data produced in the present study are available upon reasonable request to the authors

## Acknowledgements

The authors would like to thank individuals with Long Covid and healthcare professionals who provided valuable suggestions and feedback during the iterative process of scale development

## Using the scale

The Modified C19-YRS paper version is free to use, and a copy of the tool is available as Appendix of this paper and also on the University of Leeds website. The digital system developed by ELAROS comprises of a smartphone application for the patient and a web portal for the clinicians managing the care of the patient. Any clinical service wishing to acquire the system can contact ELAROS who will provide a demonstration of the system and provide necessary training to the users of the system.

## Notes

### Competing Interest Statement

The authors have declared no competing interest.

### Funding Statement

This work is supported by University of Leeds Medical Research Council (MRC) Confidence in Concept (CiC) grant for psychometric evaluation of C19-YRS.

### Author Declarations

University of Leeds School of Medicine Research Ethics Committee

