## Supplementary material for "The modified COVID-19 Yorkshire Rehabilitation Scale (C19-YRSm) patient-reported outcome measure for Long Covid or Post-COVID syndrome": Modified C19-YRS

### Modified COVID-19 Yorkshire Rehabilitation Screening (C19-YRS)

#### Self-report version

Patient name:

Hospital number:

Date: Time:

*The purpose of this questionnaire is to find out more about your current problems following COVID-19 illness. Your responses will be recorded in your clinical notes. We will use this information to monitor your symptoms, offer treatments and assess response to treatment.*

*This questionnaire will take around 15 minutes. If there are any topics you don't want to talk about you can choose not to respond.*

*Do you consent for this information to be used for audit and research as well ?* **Yes** ☐ **No** ☐

**SYMPTOM SEVERITY**

*Please answer the questions below to the best of your knowledge.*
***'Now'*** *refers to how you feel now/this week (last 7 days).*
***"Pre-COVID"*** *refers to how you were feeling prior to contracting the illness.*
*If you are unable to recall this, just state 'don't know'*

*Rate the severity of each problem on a scale of 0-3:*
***0 = None; no problem***
***1 = Mild problem; does not affect daily life***
***2 = Moderate problem; affects daily life to a certain extent***
***3 = Severe problem; affects all aspects of daily life; life-disturbing***

| | | | |
|---|---|---|---|
| 1. Breathlessness | Breathlessness: | **Now** | **Pre-COVID** |
| | a) At rest | **0** ☐ **1** ☐ **2** ☐ **3** ☐ | **0** ☐ **1** ☐ **2** ☐ **3** ☐ |
| | b) Changing position e.g. from lying to sitting or sitting to lying | **0** ☐ **1** ☐ **2** ☐ **3** ☐ | **0** ☐ **1** ☐ **2** ☐ **3** ☐ |
| | c) On dressing yourself | **0** ☐ **1** ☐ **2** ☐ **3** ☐ | **0** ☐ **1** ☐ **2** ☐ **3** ☐ |
| | d) On walking up a flight of stairs | **0** ☐ **1** ☐ **2** ☐ **3** ☐ | **0** ☐ **1** ☐ **2** ☐ **3** ☐ |
| 2. Cough/ throat sensitivity/ voice change | Cough/ throat sensitivity | **0** ☐ **1** ☐ **2** ☐ **3** ☐ | **0** ☐ **1** ☐ **2** ☐ **3** ☐ |
| | Change of voice | **0** ☐ **1** ☐ **2** ☐ **3** ☐ | **0** ☐ **1** ☐ **2** ☐ **3** ☐ |

| | | | |
|---|---|---|---|
| 3. Fatigue (tiredness not improved by rest) | Fatigue levels in your usual activities | **0** ☐ **1** ☐ **2** ☐ **3** ☐ | **0** ☐ **1** ☐ **2** ☐ **3** ☐ |
| 4. Smell/taste | Altered smell | **0** ☐ **1** ☐ **2** ☐ **3** ☐ | **0** ☐ **1** ☐ **2** ☐ **3** ☐ |
| | Altered taste | **0** ☐ **1** ☐ **2** ☐ **3** ☐ | **0** ☐ **1** ☐ **2** ☐ **3** ☐ |
| 5. Pain/discomfort | Chest pain | **0** ☐ **1** ☐ **2** ☐ **3** ☐ | **0** ☐ **1** ☐ **2** ☐ **3** ☐ |
| | Joint pain | **0** ☐ **1** ☐ **2** ☐ **3** ☐ | **0** ☐ **1** ☐ **2** ☐ **3** ☐ |
| | Muscle pain | **0** ☐ **1** ☐ **2** ☐ **3** ☐ | **0** ☐ **1** ☐ **2** ☐ **3** ☐ |
| | Headache | **0** ☐ **1** ☐ **2** ☐ **3** ☐ | **0** ☐ **1** ☐ **2** ☐ **3** ☐ |
| | Abdominal pain | **0** ☐ **1** ☐ **2** ☐ **3** ☐ | **0** ☐ **1** ☐ **2** ☐ **3** ☐ |
| 6. Cognition | Problems with concentration | **0** ☐ **1** ☐ **2** ☐ **3** ☐ | **0** ☐ **1** ☐ **2** ☐ **3** ☐ |
| | Problems with memory | **0** ☐ **1** ☐ **2** ☐ **3** ☐ | **0** ☐ **1** ☐ **2** ☐ **3** ☐ |
| | Problems with planning | **0** ☐ **1** ☐ **2** ☐ **3** ☐ | **0** ☐ **1** ☐ **2** ☐ **3** ☐ |
| 7. Palpitations/ dizziness | Palpitations in certain positions, activity or at rest | **0** ☐ **1** ☐ **2** ☐ **3** ☐ | **0** ☐ **1** ☐ **2** ☐ **3** ☐ |
| | Dizziness in certain positions, activity or at rest | **0** ☐ **1** ☐ **2** ☐ **3** ☐ | **0** ☐ **1** ☐ **2** ☐ **3** ☐ |
| 8. Post-exertional malaise (worsening of symptoms) | Crashing or relapse hours or days after physical, cognitive or emotional exertion | **0** ☐ **1** ☐ **2** ☐ **3** ☐ | **0** ☐ **1** ☐ **2** ☐ **3** ☐ |
| 9. Anxiety/ mood | Feeling anxious | **0** ☐ **1** ☐ **2** ☐ **3** ☐ | **0** ☐ **1** ☐ **2** ☐ **3** ☐ |
| | Feeling depressed | **0** ☐ **1** ☐ **2** ☐ **3** ☐ | **0** ☐ **1** ☐ **2** ☐ **3** ☐ |
| | Having unwanted memories of your illness or time in hospital | **0** ☐ **1** ☐ **2** ☐ **3** ☐ | **0** ☐ **1** ☐ **2** ☐ **3** ☐ |
| | Having unpleasant dreams about your illness or time in hospital | **0** ☐ **1** ☐ **2** ☐ **3** ☐ | **0** ☐ **1** ☐ **2** ☐ **3** ☐ |
| | Trying to avoid thoughts or feelings about your illness or time in hospital | **0** ☐ **1** ☐ **2** ☐ **3** ☐ | **0** ☐ **1** ☐ **2** ☐ **3** ☐ |
| 10. Sleep | Sleep problems, such as difficulty falling asleep, staying asleep or oversleeping | **0** ☐ **1** ☐ **2** ☐ **3** ☐ | **0** ☐ **1** ☐ **2** ☐ **3** ☐ |

**FUNCTIONAL ABILITY**

| | | | |
|---|---|---|---|
| 11. Communication | Difficulty with communication/word finding difficulty/understanding others | **Now** | **Pre-COVID** |
| | | **0** ☐ **1** ☐ **2** ☐ **3** ☐ | **0** ☐ **1** ☐ **2** ☐ **3** ☐ |
| 12. Walking or moving around | Difficulties with walking or moving around | **0** ☐ **1** ☐ **2** ☐ **3** ☐ | **0** ☐ **1** ☐ **2** ☐ **3** ☐ |
| 13. Personal care | Difficulties with personal tasks such as using the toilet or getting washed and dressed | **0** ☐ **1** ☐ **2** ☐ **3** ☐ | **0** ☐ **1** ☐ **2** ☐ **3** ☐ |
| 14. Other activities of Daily Living | Difficulty doing wider activities, such as household work, leisure/sporting activities, paid/unpaid work, study or shopping | **0** ☐ **1** ☐ **2** ☐ **3** ☐ | **0** ☐ **1** ☐ **2** ☐ **3** ☐ |
| 15. Social role | Problems with socialising/interacting with friends* or caring for dependants<br><br>*related to your illness and not due to social distancing/lockdown measures | **0** ☐ **1** ☐ **2** ☐ **3** ☐ | **0** ☐ **1** ☐ **2** ☐ **3** ☐ |

**OTHER SYMPTOMS**

Please select any of the following symptoms you have experienced since your illness in the last 7 days.
Please also select any previous problems that have worsened for you following your illness.

☐ Fever
☐ Skin rash/ discolouration of skin
☐ New allergy such as medication, food etc
☐ Hair loss
☐ Skin sensation (numbness/tingling/itching/nerve pain)
☐ Dry eyes/ redness of eyes
☐ Swelling of feet/ swelling of hands
☐ Easy bruising/ bleeding
☐ Visual changes
☐ Difficulty swallowing solids
☐ Difficulty swallowing liquids
☐ Balance problems or falls
☐ Weakness or movement problems or coordination problems in limbs
☐ Tinnitus
☐ Nausea
☐ Dry mouth/mouth ulcers
☐ Acid Reflux/heartburn
☐ Change in appetite
☐ Unintentional weight loss
☐ Unintentional weight gain
☐ Bladder frequency,urgency or incontinence

☐ Constipation, diarrhoea or bowel incontinence
☐ Change in menstrual cycles or flow
☐ Waking up at night gasping for air (also called sleep apnea)
☐ Thoughts about harming yourself

Other symptoms – free text

**OVERALL HEALTH**

How good or bad is your health overall in the last 7 days?

For this question, a score of 10 means the BEST health you can imagine. 0 means the WORST health you can imagine.

a) Now: WORST HEALTH 0 ☐ 1 ☐ 2 ☐ 3 ☐ 4 ☐ 5 ☐ 6 ☐ 7 ☐ 8 ☐ 9 ☐ 10 ☐ BEST HEALTH
b) Pre-Covid: WORST HEALTH 0 ☐ 1 ☐ 2 ☐ 3 ☐ 4 ☐ 5 ☐ 6 ☐ 7 ☐ 8 ☐ 9 ☐ 10 ☐ BEST HEALTH

**EMPLOYMENT**

Occupation: _______________________

Has your COVID-19 illness affected your work?

☐ No change
☐ On reduced working hours
☐ On sickness leave
☐ Changes made to role/ working arrangements (such as working from home or lighter duties)
☐ Had to retire/ change job
☐ Lost job

Any other comments/concerns:________________________________________________

**PARTNER/FAMILY/CARER PERSPECTIVE**

This is space for your partner, family or carer to add anything from their perspective:
